# Self-Reliance, Social Norms, and Self-Stigma as Barriers to Psychosocial Help Seeking Among Rural Cancer Survivors with Cancer-Related Distress

**DOI:** 10.1101/2021.07.13.21260150

**Authors:** Pamela B. DeGuzman, David L. Vogel, Veronica Bernacchi, Margaret A. Scudder, Mark J. Jameson

**Author notes:** **Corresponding author:** Please address all correspondence to Pamela B. DeGuzman; University of Virginia School of Nursing; PO Box 800782; Charlottesville, VA. 22902. **Funding:** This study was funded by a grant from the University of Virginia Cancer Center.

## Abstract

**Objectives:** Even when technology allows rural cancer survivors to connect with supportive care providers from a distance, uptake of psychosocial referrals is low. During our telemedicine-delivered intervention aimed at identifying rural survivors with high distress and connecting them with psychosocial care, fewer than 1/3 of those with high distress accepted a referral. The purpose of this research was to examine the reasons rural cancer survivors did not accept a psychosocial referral.

**Methods:** We utilized a qualitative descriptive design to analyze data from interviews conducted with participants who had been offered a psychosocial referral during the intervention. Interviews were conducted 6 weeks following the intervention (n=14) and 9 months after the completion of the intervention (n=6).

**Results:** Ultimately, none of the rural cancer survivors in our study engaged with a psychosocial care provider, including those who had originally accepted a referral for further psychosocial care. When explaining their decisions, survivors minimized their distress, emphasizing their self-reliance and the need to handle distress on their own. They expressed a preference for dealing with distress via informal support networks, which was often limited to close family members. No survivors endorsed public stigma as a barrier to accepting psychosocial help, but several suggested that self-stigma associated with not being able to handle their own distress was a reason for not seeking care.

**Significance of Results:** Rural cancer survivors’ willingness to accept a psychosocial referral may be mediated by the rural cultural norm of self-reliance, and by self-stigma. Interventions to address referral uptake may benefit from further illumination of these relationships as well as a strength-based approach that emphasizes positive aspects of the rural community and individual self-affirmation.

## Introduction

Cancer survivors from rural areas experience high-levels of cancer-related distress (CRD; the multifactorial, unpleasant, emotional experience that interferes with their ability to cope with cancer, treatment, and symptoms effectively). As recently as 10 years ago, fewer than 10% of all individuals with cancer were being screened for CRD, but with wider adoption of screening tools, such as the National Comprehensive Cancer Network Distress Thermometer and Problem List (Holland et al., 2013), individuals with cancer are now being screened at higher rates, above 70% in some cases (Carlson et al., 2020; Skaczkowski et al., 2018, 2020). Unfortunately, despite this high rate of screening, difficultly connecting rural survivors with psychosocial support persists. In fact, rates of successful referral of rural survivors to psychosocial services remains low (Bauwens et al., 2014; Funk et al., 2016; Opie et al., 2017), leaving rural survivors highly vulnerable to a range of negative sequelae of unmet needs including a higher risk of suicide (DeGuzman et al., 2017; Deleemans et al., 2020; Hubbard et al., 2015; Tzelepis et al., 2018).

Distance to care is well-accepted as a significant factor in rural survivors’ reluctance to receive referral for further psychosocial care (Andrykowski & Burris, 2010; Opie et al., 2017; Pesut et al., 2010). Yet, it fails to fully explain survivors’ unwillingness to receive psychosocial care when distance barriers can be successfully be overcome via technology. For example, we conducted a telemedicine-delivered intervention with rural cancer survivors to screen for CRD and make referrals for lingering post-treatment unmet needs. The intervention, Comprehensive Assistance: Rural, Nursing and Guidance (CARING) was designed to overcome technology barriers experienced by many living in broadband-poor areas, offering options for survivors without home-based internet access to connect to an oncology-specialized nurse (DeGuzman et al., 2020, 2021). Participants (*n* = 14) who were found to have high CRD (through a combination of self-identification and nursing assessment) related to a psychosocial issue were referred for psychosocial support (e.g., oncology-specialized social worker [SW] from the Cancer Center). SWs at the Cancer Center where the study took place initially contact patients by phone to discuss the range of support options available, including telephone counseling and finding resources in their area such as support groups. However, despite this direct contact, fewer than one-third (i.e., 28.6%) of the 14 accepted the referral for further psychosocial care (DeGuzman et al., 2020, 2021).

Given this low acceptance of psychosocial services, even when distance barriers were removed, there is need to better understand the psychological reasons why referrals are rejected in order to better develop interventions to increase acceptance. However, only one study has directly examined rural cancer survivors’ reasons for not seeking psychosocial services. In this study, rural cancer survivors reported that speaking with a psychologist or using a support group to deal with psychosocial issues is not an accepted social norm, suggesting that relying on those within their own personal circle is more acceptable than utilizing professional care to deal with non-physical issues that arise from cancer treatment (Andrykowski & Burris, 2010). Other studies on help seeking in rural settings have also noted the high value that is placed on self-reliance and privacy (Fennell et al., 2018). Men in the study also reported a desire to minimize or normalize the problem, and to have emotional control. Research has also suggested that stigma may be a barrier within rural communities, as seeking help is viewed as a sign of personal weakness (Fuller et al., 2000), and that rural women are afraid to ask for help with mental health issues because of stigma (Snell-Rood et al., 2017). Specifically, higher rates of perceived public stigma (i.e. the perception that others view those who seek help for mental health concerns as “weak” or “crazy”; Vogel et al., 2009) and self-stigma (i.e., the perception of oneself as being inferior or a failure if seeking help with a mental health concern; Vogel et al., 2006) have been shown to exist in rural populations (Schroeder et al., 2021; Stewart et al., 2015). However, despite these important findings, this past research is largely limited to individuals not currently experiencing distress or who are asked to respond to a hypothetical situation. Research has not focused on individuals currently experiencing distress Swho are offered services. Given our research with rural cancer survivors, where we found that only 28.6% of rural survivors with high CRD accepted a referral for psychosocial care (DeGuzman et al., 2021), the purpose of this research was to directly examine the reasons rural cancer survivors did not accept a psychosocial referral.

## Methods

We utilized a qualitative descriptive design to accomplish the study purpose (Sandelowski, 2000). Six weeks following the CARING study, post-intervention interviews were originally conducted with participants. For the present analysis, we evaluated data from the interviews of the 14 participants to whom the nurse had offered a psychosocial referral. Of the 14, four had accepted the referral and 10 had declined (DeGuzman et al., 2021). The 10 who declined a referral during the intervention were asked about their reasons for declining. The 4 participants who had accepted the referral during the intervention were asked if they had yet heard from the referring provider and if the process had moved forward. For the current study, we analyzed data from the 14 original post-intervention interviews, and attempted to re-contact each participant for a second interview 9 months after the conclusion of the intervention, targeting a more in-depth understanding of their perspectives on the barriers to their acceptance of a psychosocial referral.

Interview questions for this second interview were drawn from validated instruments designed to understand individual’s reasons for not accepting or following through on the referral. Specifically, participants were asked if their decision to accept or not accept a referral was related to self-reliance (i.e., feelings of not being able to take care of one’s own problems; Fennell et al., 2018); public stigma related to seeking mental health [i.e., (a) “others viewing them negatively or in a less favorable light”; (b) “others thinking bad things about them”; or (c) “others seeing them as seriously disturbed or thinking they posed a risk to others”]; and self-stigma related to seeking mental health [i.e., if accepting a psychosocial referral would impact them feeling (a) “inadequate,” (b) “inferior,” (c) or “less satisfied with themselves”] (Vogel et al., 2006, 2009). For the 4 participants who had originally accepted a referral, we also sought to determine if they had met with the Cancer Center SW or other psychosocial care providers.

We used a deductive, iterative approach to coding the data, guided by qualitative content methods (Sandelowski, 2000). We categorized data into known barriers to mental health care in rural populations. Categories were developed into themes; categories and themes were discussed among members of the research team to obtain consensus. The Institutional Review Board for Health Research at the University of Virginia approved the study. Verbal consent was obtained from all participants.

## Results

In addition to analyzing the data from the 14 post-intervention interviews and were able to successfully re-contact 6 of the original 14 for long-term follow-up. Two had passed away since the intervention and we were unable to reach the remaining 6. Of the 6 survivors that we recontacted, 2 of the female participants had originally accepted a referral for an oncology-specialized social worker from the Cancer Center, but when the SW contacted them, neither had followed through with receiving further assistance.

**Table 1** presents demographics for the 14 participants. The sample was equally split among male and female, with an average age of 62. The majority were white, non-Hispanic, and the most common cancer was squamous cell carcinoma of the oral cavity. Over 85% of participants had cancer that had not spread to lymph nodes or metastasized elsewhere. The mean distress score was 5.8/10.

**Table 1.** Participant Characteristics (n=14)

| Characteristic | N (%) |
| --- | --- |
| Gender |  |
| Male | 7 (50.0) |
| Female | 7 (50.0) |
| Age (mean; sd; range) | 62; 12.4; 39-80 |
| Race |  |
| White | 11 (78.6) |
| Black | 2 (14.3) |
| Other | 1 (7.1) |
| Ethnicity |  |
| Non-Hispanic | 12 (85.7) |
| Hispanic | 2 (14.3) |
| Cancer Site |  |
| Oral Cavity | 5 (35.7) |
| Thyroid | 4 (28.6) |
| Pharynx | 2 (14.3) |
| Other | 2 (14.3) |
| More than one site | 1 (7.1) |
| Cancer type |  |
| Squamous Cell Carcinoma | 8 (57.1) |
| Papillary Thyroid Carcinoma | 4 (28.6) |
| Other | 2 (14.3) |
| Cancer Stage |  |
| Early | 12 (85.7) |
| Late | 2 (14.3) |
| Distress Score (mean; sd; range) | 5.8; 0.3; 5.17-7.17 |

We ultimately extracted three themes from the data: *Minimization and Self-Reliance*; *Preference for Use of Informal Support*; and *Self-Stigma for Some, but no Public Stigma*.

### Minimization and Self-Reliance

When explaining their rationale for not pursuing further psychosocial care, 6 participants made light of their distress by minimizing it, instead focusing on positivity and self-reliance. A positive attitude was viewed as a means to regain health. One male survivor stated “Yeah well that’s the only way to get better, isn’t it? …I certainly try not to ever feel sad. I feel lucky [that the cancer is in remission].” Several laughed while describing their distress. A male survivor who had declined speaking with a SW insisted that his situation was not negative, and that he could– and should–handle it himself: “I had a brain bleed. They drilled a couple of holes in my head. But life goes on! There was uh…a series of complications with the cancer. This is just a series of misfortunes the way I look at it [laughs], and I have to deal with them.” He then emphasized his self-reliance and strength by stating: “And I do deal with them… I’m not an invalid or anything.” One female survivor minimized her fear of recurrence, continuously reframing her fears of recurrence as “concerns, stating “it’s just those same concerns. You know I get this sore throat and it’s like, it’s just a concern that the cancer may come back like the other time. So, it’s just a concern that it’ll come back like the other two times… it’s not even the fear it’s just the concern of it, of it coming back.” The idea of CRD not rising to the level of requiring outside intervention was echoed by other survivors. Another female survivor who declined a referral to the Cancer Center SW stated, I just tend to view things I want to handle them myself…If I felt an absolute need [to speak to someone about my CRD] I would do it…I didn’t feel an urgent need, to be honest.”

### Preference for Use of Informal Support

Rural survivors in our study described a small circle of people to whom they would speak about their CRD. They reported limiting these discussions to close family, but occasionally included close friends. A female survivor turned down the referral to the Cancer Center SW by explaining “my niece is a social worker and we’ve had our chats.” Two male survivors reported only discussing distress with their spouses: When asked why he had not been interested in speaking with the Cancer Center SW, a male survivor replied, “No. I’ve got great support from my wife as a caregiver.” A second male survivor reported that he spoke to his wife about his CRD, but only minimally. I’m uh, I’m just not a big talker [laughs]. My wife’s always gettin’ on me.

That’s just uh, just kinda the way I am.” A female survivor expressed a similar sentiment: “What you might consider an inadequacy, or a problem or an issue, I handle them myself, and that’s just the way I am…It would take some getting beyond the point of me handling it entirely on my own and feeling comfortable in sharing with someone else.” However, she also suggested that her family was included as part of her handling things herself: “My family is close, husband is very close…those are my anchors.”

### Self-Stigma for Some, but no Public Stigma

Participants did not report public stigma associated with seeking help for CRD. One female survivor who reported high distress during the intervention but turned down a referral to the SW stated, “I don’t think anyone would say anything bad about [my speaking to a social worker].” Despite our participants’ sense that society would not judge them harshly, a few of them did indicate they would judge themselves negatively if they had followed up with a SW or other MH care provider. When a female survivor who had originally accepted a referral, but then never called the SW back was asked if speaking to the SW would have made her feel that she could not handle her problems herself, she confirmed, “I would question it…I don’t want to learn [sic] the appearance that as I’m getting older, I am less able to handle my situation, and that’s a protection on my part. I try not to view anybody else that way, but I tend to view myself more critical [sic].” Not all survivors shared this perception. An male survivor with high distress who turned down the SW referral stated that he did not believe would not have felt inadequate if he had accepted help from the SW: “I wouldn’t think there was any stigma to it.” A female survivor explained that she had not accepted a referral to the SW because, “I have a therapist.” She went on to explain her perspective about the impact of socioeconomic status on her views about seeking mental healthcare: “I make over $100,000, you know, I have a bunch of degrees and everything…but I do live in a rural area… I think for me there aren’t those barriers to treatment.”

Despite many of the survivor’s statements suggesting that seeking MH help for CRD is acceptable, overwhelmingly our participants still declined the opportunity to speak to a SW or counselor. During the intervention, a male survivor self-reported high levels of nervousness and worry related to his cancer diagnosis, treatment and the possibility of recurrence, but declined an offer for a social worker or participation in a support group. During the follow up interview he reported feeling no public or self-stigma associated with seeking help for his CRD, but was unable to articulate his reasons for refusing, stating “The social worker thing was something that I did not feel would help me.”

## Discussion

Despite high levels of CRD and recommendations from an oncology-specialized nurse, none of the participants in our study – not even those who had originally agreed to accept a referral—ultimately used the Cancer Center’s psychosocial resources. When explaining their views toward not accepting help, survivors tended to minimize their experiences of distress while emphasizing self-reliance and a desire to only speak to close family and occasionally friends. Research has found similar phenomena in rural populations. For example, previous work with cancer survivors has shown a desire to relying on family and friends to deal with psychosocial issues (Andrykowski & Burris, 2010). Several studies have also found self-reliance and problem-minimization as barriers to seeking mental health treatment for rural populations (Fennell et al., 2018; Judd et al., 2006; Komiti et al., 2006; Snell-Rood et al., 2017). Similarly, a qualitative study with psychosocial care providers serving a rural Australian region, reported that participants noted that the distress of residents of their region had to rise to a very serious threshold before residents would even acknowledge its existence (Fuller et al., 2000). Taken together with our results, this suggests that the social norms of self-reliance and desire to not share personal information with someone from outside the survivors’ immediate circle, may be key contributing factors, as well as important intervention points to address, to not seeking help for psychological distress among rural cancer survivors. This might be particularly important, as prior research indicates that limiting mental healthcare to discussions with family and friends is insufficient to address the profound distress that survivors experience, and that those who elect to seek professional care find it highly impactful (Gunn et al., 2013).

While consistent with past research (Fennell et al., 2018), both male and female survivors in our study emphasized positive thinking, self-reliance, and minimization of distress, our findings also suggested some potential differences in the approach to addressing barriers to care for men and women. Specifically, men in our study reported a smaller circle of trust, which typically just included their wives. Consistent with this, a study of 409 rural Australian men and women found that rural men reported more barriers to seeking mental health care than women. Other work has also found that certain norms such as a desire for stoicism and emotional control to be a stronger barrier to help-seeking for rural men than women (Fennell et al., 2018; Judd et al., 2006). As such, since rural women are more likely to seek mental healthcare than rural men (Judd et al., 2006; Wrigley et al., 2005), approaches to overcome barriers in rural male cancer survivors may benefit from examining gender-focused interventions. For example, our results may suggest that a dyadic intervention that includes their spouse or caregiver may be particularly salient for men.

The current results are consistent with some assertions that stigma is a moderately important barrier to help seeking, reported by approximately a quarter to one-third of participants across multiple research studies (Clement et al., 2015). Interestingly, while stigma is the most cited barrier to seeking help, rural cancer survivors in our study did not report public stigma associated with seeking help for CRD as a barrier to accepting a referral. None of the six individuals we re-contacted reported that others would view them negatively if they were to seek help from the Cancer Center SW. In turn, self-stigma associated seeking psychosocial services was found to be a limiting factor from accepting a psychosocial referral, at least for some participants. One possible explanation for this is that the cancer experience is viewed by others as sufficiently debilitating physically and emotionally, that it is considered warranting emotional support for others, but still does not rise to the level of warranting breaking social norms oneself. One of our participants referred specifically to this, stating that she tried not to view others that way, but viewed herself more critically. This finding is also consistent with most research showing that self-stigma is a more salient barrier to seeking help than public stigma in general populations (Vogel et al., 2007) and with the research on rural communities regarding the presence of self-stigma (i.e., seeking help is viewed as a sign of personal weakness; Fuller et al., 2000; Vogel et al., 2006). As such, self-stigma may be an important barrier for those who endorse it, and not salient for everyone.

Alternatively, it may be that stigma is an important factor that influences other factors and thus its systemic effects may not always be noticeable (Clement et al., 2012). Others have also suggested that stigma is part of interrelated network of barriers (Schomerus & Angermeyer, 2008). For example, Jennings and colleagues examined a model in which public stigma was linked to attitudes towards seeking professional services through self-stigma and then self-reliance (Jennings et al., 2015). In other words, self-stigma and self-reliance were mediators and thus more proximal to decisions to seek help (and thus may be more accessible to participants’ awareness). This finding is consistent with the current findings where potential mediating factor such as self-reliance and desire to only disclose to close others were widely endorsed and stigma factors less so. Thus, similar to Jennings et al., we encourage future researchers and clinicians continue to examine the complex relationships between the different types of stigma and other factors such as self-reliance in order to be able to develop more focused interventions to increase use of services by those who could benefit.

### Implications for Cancer Survivorship Care

These findings provide a strong direction for developing interventions aimed at improving access to psychosocial care for rural cancer survivors. Barriers to rural access are typically grouped into four domains: people, place, provider, and payment (MacKinney et al., 2014). In light of increased levels of insurance coverage (Newkirk II & Damico, 2014), and ongoing technological advances for overcoming distance-related barriers to care (DeGuzman et al., 2020), personal and cultural belief systems need to be explored further. Thus, to continue enhancing equitable access, a more rigorous understanding of how rural cultural belief systems are limiting survivors’ openness to high quality, psychosocial is needed. Instruments are available that differentiate barriers (Brenner et al., 2020; Vogel et al., 2006, 2009); however, customization to illuminate rural-specific barriers may be needed. As such, researchers may consider amending instruments with questions specific to the rural populations (i.e., self-reliance, social norms around who to self-disclose, self-stigma).

Keeping in mind these barriers, clinicians caring for rural survivors may need to be aware of any negative perceptions of rural culture that are embedded in an urban bias. Current approaches toward improving access to care have been developed in the context of an urban health care delivery system (Mohatt & Mohatt, 2020). Access to mental health care in rural communities does suffer from limited access which may be further confounded by other factors, but rather than viewing aspects of rural culture through a deficit lens (i.e., stigma and self-reliance as barriers to access), we should strive to develop culturally-appropriate interventions that leverage the considerable strengths of the rural setting (i.e., resilience, strong community networks) to design effective interventions that connect rural survivors with care (Bernacchi et al., 2021).

One possible strength-based approach, that might be particularly salient to address this, is based in self-affirmation theory (Sherman & Cohen, 2006; Sherman & Hartson, 2011; Steele, 1988). Self-affirmation theory notes that we are inherently motivated to keep a positive sense of self-worth, and when we experience information that could decrease positive self-perceptions (i.e., reduce the belief that we are self-reliant and self-sufficient), we are driven to protect these positive views of the self, leading to avoidance of treatment for mental health issues (Lannin et al., 2013; Seidman et al., 2019). Fortunately, self-affirmation theory also asserts that we can reduce this drive to protect our self-worth, and thus increase likelihood of seeking therapy, by using self-affirmations (i.e., reflecting on a positive and self-relevant personal characteristic or values) to increasing self-worth prior to confronting information that could decrease these positive self-perceptions. Self-affirmation interventions, have started to receive some support in increasing use of psychosocial services (Lannin et al., 2013; Seidman et al., 2018, 2019), but needs to be further evaluated with rural cancer survivors, as this approach, if tailored to this population, may be able to directly reduce stigma and self-reliance as barriers to access.

### Study Limitations

Our study was qualitative and as such, findings were meant to provide a direction for further research and not meant to be generalizable. Still, there are several limitations to our work that may impact our findings. Our sample size of 14 was small, and we were only able to recontact 6 to discuss specific barriers. It is possible that with a larger sample size we may have found additional themes not identified in this study. The number of participants also made it difficult to fully evaluate differences in perceptions between rural women and men. Finally, we did not collect data on participants’ educational level, which is a significant driver of differences in stigma perceptions, and was highlighted by one of our participants. More precise information about barriers will be important to developing customized interventions for rural individuals and should incorporate these data.

### Conclusion

This study is the first to highlight the complex interplay of self-reliance, rural social norms, and self-stigma as a barrier to connecting rural survivors with psychosocial care. Further, unlike past stigma research, our research focuses on individuals currently experiencing distress who were making an active decision to seek help or not. Our findings suggest a strength-based approach that acknowledges positive aspects of rural communities (i.e., resilience, community trust) as well as individual strengths are components that should be incorporated into interventions to increase psychosocial referral uptake in this population.

## Data Availability

Data is not available due to participant privacy

## Disclosures and Acknowledgements

The authors have no conflicts of interests to disclose. This study was supported by a grant from the University of Virginia Cancer Center.

